# Chloroquine Administration in Breastfeeding Mothers Associates with Increased HIV-1 Plasma Viral Loads

**DOI:** 10.1101/2020.04.29.20085308

**Authors:** William A Paxton, Marloes A Naarding, Ferdinand WNM Wit, Nienke J Veldhuijzen, Matthew F Chersich, Brigitte Kankindi, Rene A Douma, Samuel Tuyizere, Suzanne Jurriaans, Rolf W Sparidans, Jos H Beijnen, Georgios Pollakis, Johan R Boelaert, Joep MA Lange, Joseph Vyankandondera, Stanley Luchters

**Affiliations:** Department of Clinical Infection, Microbiology and Immunology, Institute of Infection, Veterinary and Ecological Sciences, University of Liverpool, Liverpool, UK; Laboratory of Experimental Virology, Department of Medical Microbiology, Amsterdam UMC, Locat AMC, University of Amsterdam, Amsterdam, Netherlands; Amsterdam Institute for Global Health and Development, Amsterdam, Netherlands; Univ Witwatersrand, Fac Hlth Sci, Wits Reprod Hlth & HIV Inst, Johannesburg, South Africa; CHUK, Centre Hospitalier Universitaire de Kigali and Belgian Technical Cooperation, Kigali, Rwanda; Laboratory of Clinical Virology, Department of Medical Microbiology, Amsterdam UMC, Locat AMC, University of Amsterdam, Amsterdam, Netherlands; Universiteit Utrecht, Department of Pharmaceutical Sciences, Section of Biomedical Analysis, Division of Drug Toxicology, Utrecht, The Netherlands; Internal Medicine, Unit of Renal and Infectious Diseases, Algemeen Ziekenhuis St-Jan, Brugge, Belgium; Department of Population Health, Aga Khan University, Nairobi, Kenya; Department of Public Health and Primary Care, International Centre for Reproductive Health (ICRH), Ghent University, Ghent, Belgium; Department of Epidemiology and Preventive Medicine, Monash University, Australia; Burnet Institute, Australia

**Keywords:** HIV-1, CQ, HCQ, post-partum, randomized placebo-controlled trial

## Abstract

Chloroquine (CQ) and Hydroxychloroquine (HCQ) have been proposed to be effective at treating COVID-19 patients. We, and others, have previously reported on the capacity of CQ to reduce HIV-1 replication *in vitro*. We tested CQ administration in post-partum mothers on influencing HIV-1 viral loads in human milk as a means of lowering mother to child transmission. A Phase I/II, randomized, placebo-controlled study to evaluate chloroquine administration to reduce HIV-1 RNA levels in human milk: the CHARGE study. Thirty HIV-1 positive pregnant Rwandese women (CQ *n* = 20; placebo *n* = 10) were enrolled in a 16-week study, with the treatment group receiving a 200 mg oral dose of CQ daily. Base-line plasma viral load (pVL) measurements and CD4 counts were determined prior to delivery, and pVL, breast milk VL (bmVL) and CQ levels measured during treatment. For women receiving treatment, CQ concentration was higher in breast milk compared to plasma (over 2.5-fold), with a positive correlation between the levels in the two compartments (P < 0.003). A link between high CQ concentrations in plasma and high CD4 counts (*P* < 0.001) was observed. Surprisingly, we found a significant increase in pVL after CQ treatment in over half of the mothers (n=11; *P* < 0.001) and with no alteration to bmVL measurements. No specific amino acid alterations in the gp120 envelope sequences could be associated with CQ administration. CQ usage is associated with a significant increase to pVL in early breastfeeding mothers from Rwanda which cautions against the use of CQ in such individuals. Our results highlight a discrepancy between CQ effects on modulating HIV-1 replication *in vitro* versus *in vivo* and indicate caution when prescribing CQ to postpartum HIV-1 untreated mothers. This discrepancy should be taken into consideration when testing CQ or HCQ treatment in COVID-19 clinical trials, especially relating to the post-partum setting.

## Introduction

The extent of COVID-19 disease (SARS-CoV-2) has accelerated efforts to identify readily-available, safe and effective or vaccines. Given its antiviral properties, studies are investigating the effectiveness of chloroquine (CQ) and hydroxychloroquine (HCQ) to treat people with confirmed coronavirus infection.

Chloroquine and HCQ are cheap, widely available and have been shown to reduce HIV-1 replication *in vitro* and *in vivo* [1–5]. We have previously shown that CQ can interfere with virus replication in CD4^+^ T lymphocytes and that the inhibition is likely conferred at the cellular level [6]. It has been shown that CQ treatment can interfere with the potential N-linked glycosylation (PNG) pattern of the gp120 envelope protein with the reported modulation of the 2G12 monoclonal antibody epitope [1] and the capacity by which HIV-1 can undergo viral transfer to CD4^+^ T lymphocytes via cells expressing DC-SIGN *in vitro* [6]. The endosomal pH increases with CQ treatment and is believed to reduce IL-6 production [7], thereby causing a down-regulation of HIV-1 production in chronically infected T-cells and monocyte cell lines [2,8]. Several clinical trials have previously shown that CQ and HCQ treatment can reduce pVL *in vivo* with no effect on CD4 counts [3,4].

Mother to child transmission of HIV-1 (MTCT) can occur either *in-utero*, *intra-partum* or via breastfeeding [9–11]. Transmission of HIV-1 to the infant via breastfeeding has been associated with high pVL and breast milk VL (bmVL), low CD4 count and the presence of sub-clinical mastitis [13,14]. CQ has been shown to be secreted in human milk [15–17] suggesting it may be effective at lowering bmVL. The presence of CQ in human milk has also been shown to be non-toxic to children [18], making it ideally suitable for such a purpose. This study was undertaken to test whether CQ treatment of HIV-1 infected breastfeeding mothers could be effective at lowering VL and potentially limiting MTCT, and may provide valuable lessons for its effect with other viral infections, such as SARS-CoV-2.

## Methods

### Study design, Participants and clinical evaluations

Between 2000–2001, we conducted a phase I/II randomized placebo-controlled pilot trial undertaken in Rwanda with 30 HIV-1 breastfeeding mothers randomized (2:1) to receive oral CQ 200 mg once daily or placebo (Pl) for 16 weeks (wks). All women received single-dose nevirapine (200 mg) at start of labor and children received single-dose nevirapine within 72 hrs. The study was approved by all the appropriate Ethic Committees (IEC) in both the Netherlands, the STEG-METC (ref no R01-089) and Rwanda and informed consent was obtained for all mothers. The CQ capsules, and matched placebo capsules, were prepared at the pharmacy department of the Slotervaart Hospital, Amsterdam, and concealed to the investigators I Rwanda (double-blinded). Patients were randomized on day 1 of breastfeeding (wk0) and the study un-blinded at wk18. All participants were confirmed HIV-1 infected (rapid tests), at least 18 years of age, therapy-naive, between 32–35 wks gestation and had not received CQ in the six wks prior. Pre-screen (psc) with follow-up and counseling visits were scheduled at wk0 (delivery), wk4, 8, 16, 17 and 18. All pVL were assessed at psc, wk0, 8 and 16 and bmVL at wk0, 8 and 16 and in both breasts. CQ concentrations were measured in blood plasma and from right (RB) and left (LB) breasts at wk0, 8 and 16. Both pVL and bmVL measurements were determined using the Roche diagnostics HIV-1 RNA PCR quantification assay.

### Measuring CQ concentrations

To 1.0 ml plasma (supplemented with CQ free plasma if required) in a glass conical tube, 25 µl of internal standard solution (50 µg/ml diaminonaphthalene in methanol), 1 ml of 1M sodium hydroxide and 6 ml diethylether were added. After shaking for 10 min, the mixture was centrifuged for 5 min at ca. 2×10^3^ *g*. After storage of the tube for 1 h at −30°C, the organic layer was poured into a second tube and evaporated at 40°C under a gentle stream of nitrogen. Finally, the residue was reconstituted in 200 µl eluent using vortex-mixing and 50 µl was injected onto the HPLC. To 1.0 ml milk (supplemented with drug-free cow milk if required) in a glass conical tube, 6 ml *n*-hexane was added. The tube was shaken for 10 min and centrifuged for 5 min at ca. 2×10^3^ *g*. After storage of the tube for 1 h at −30°C, the organic layer was discarded. After thawing the aqueous layer, 1 ml of 1M sodium hydroxide and 6 ml diethylether were added and the procedure was continued according to the pre-treatment of plasma samples. Chromatographic conditions: Fifty µl injections were made on a Symmetry C_18_ column (100×4.6 mm, d_p_ = 3.5 µm, average pore diameter = 10 nm, Waters), protected by a Symmetry C_18_ pre-column (20×3.8 mm, d_p_ = 5 µm, Waters). The column temperature was maintained at ambient temperature (23–29°C). The eluent comprised 9% (v/v) acetonitrile and 91% (v/v) phosphate buffer (45 mM, pH 3.0) and the eluent flow rate was 1.0 mL/min. The UV detection wavelength was 340 nm.

### PCR amplification and sequencing of gp120

Viral RNA was isolated from serum and the C1C4 gp120 region amplified by RT-PCR as previously described with the appropriate primers [19]. PCR products were cloned into the TOPO II vector (Invitrogen) and sequenced bi-directionally using the BigDye Terminator Cycle Sequencing kit (ABI). Primers A1360 GAG CCA ATT CCY ATA CAT TAT TG, SP6 ATT TAG GTG ACA CTA TAG and T7 TAA TAC GAC TCA CTA TAG GG were used for sequencing. Subtype determination and phylogenetic analysis was performed with the neighborhood-joining (N-J) analysis of MEGA [20]. PNG sites were determined using Glycosite (http://hiv-web.lanl.gov/content/hivdb/GLYCOSITE/glycosite.html).

### Statistical analysis

Descriptive statistics for group comparison and regression analyses were performed utilizing the Prism package software (GraphPad software Inc., San Diego, California, USA, version 4.0). All available CD4 T cell counts, pVL and CQ level data were utilized, with the pVL values Log_10_-transformed. The Mann Whitney or the Wilcoxon signed rank tests were applied appropriately and the level of significance was set at *P* = 0.05.

## Results

### Clinical parameters of study individuals

The study participants are described in Table 1 (Placebo: n = 10, CQ: n = 20) and with a study schematic shown (Fig. 1). Of the 30 study individuals enrolled 7 did not finish due to early termination of breastfeeding or death of the child (Placebo: n = 5, CQ: n = 2). Infant pVL measurements taken at wk16 identify 6 infants to be infected, 1/5 in the Placebo group and 5/18 in the CQ group. From gp120 V3 region PCR analysis at wk0 we identified 3 infants to be HIV-1 positive at time of delivery with the remaining 3 infected either *peri-partum* or during the early breastfeeding period. Interestingly, the highest wk16 pVL were in those HIV-1 positive at time of delivery.

**Table 1.** Study individuals.

| Code | Gp | Subtype | Inf | pVL<br>wk8/0 | pVL<br>wk16/0 | Code | Gp | Subtype | Inf | pVL<br>wk8/0 | pVL<br>wk16/0 |
| --- | --- | --- | --- | --- | --- | --- | --- | --- | --- | --- | --- |
| 060 | Pl | A* | 0 | sna | 1.3 | 020 | CQ | n/d | 0 | **44.0** | 2.3 |
| 090 | Pl | A* | 750,000<sup>+</sup> | sna | 0.8 | 030 | CQ | A | 0 | **37.6** | **10.1** |
| 100 | Pl | A | sna | 0.4 | sna | 040 | CQ | A | 0 | 1.6 | 0.1 |
| 110 | Pl | A* | 0 | sna | 4.2 | 050 | CQ | A* | 0 | **18.5** | **27.8** |
| 150 | Pl | A | 0 | 0.7 | sna | 130 | CQ | C* | 7,590 | 0.6 | 0.6 |
| 200 | Pl | A* | 0 | 0.1 | 0.3 | 160 | CQ | A* | sna | 0.8 | sna |
| 240 | Pl | n/d | 0 | 1.5 | 1.6 | 180 | CQ | C | 0 | 0.2 | 1.4 |
| 260 | Pl | A | 0 | 0.6 | sna | 190 | CQ | C* | 0 | **19.7** | **15.6** |
| 280 | Pl | A | 0 | 1.1 | sna | 210 | CQ | C | 0 | **37.5** | **52.3** |
| 400 | Pl | A | sna | 4.3 | sna | 220 | CQ | A | 0 | 1.3 | sna |
|  |  |  |  |  |  | 250 | CQ | A/C* | 269,000<sup>+</sup> | **7.2** | **5.0** |
|  |  |  |  |  |  | 270 | CQ | C | 0 | 1.8 | 2.3 |
|  |  |  |  |  |  | 290 | CQ | C* | 531,000<sup>+</sup> | **16.1** | **121.9** |
|  |  |  |  |  |  | 300 | CQ | A* | 747,000 | **13.1** | **14.3** |
|  |  |  |  |  |  | 340 | CQ | A* | 0 | 2.8 | 4.1 |
|  |  |  |  |  |  | 350 | CQ | A | 0 | **294.0** | **10.1** |
|  |  |  |  |  |  | 360 | CQ | A | 0 | 1.0 | 1.9 |
|  |  |  |  |  |  | 370 | CQ | A* | 144,000 | **5.9** | **8.6** |
|  |  |  |  |  |  | 380 | CQ | A* | 0 | **50.9** | **12.9** |
|  |  |  |  |  |  | 390 | CQ | C | 0 | 1.1 | **8.0** |

Placebo (Pl) or CQ treated (CQ) group (Gp), subtype of infection, whether the infants are infected (Inf) and their pVL at wk16. Fold increase in mothers pVL of mothers between wk8 and wk0 (wk8/0) as well as wk16 and wk0 (wk16/0).
sna – sample not available; <sup>+</sup> - baby infected at birth; \* - gp120 amplified and sequenced;
Bold is viral load fold increase greater than 5.0.

**Table 2.**
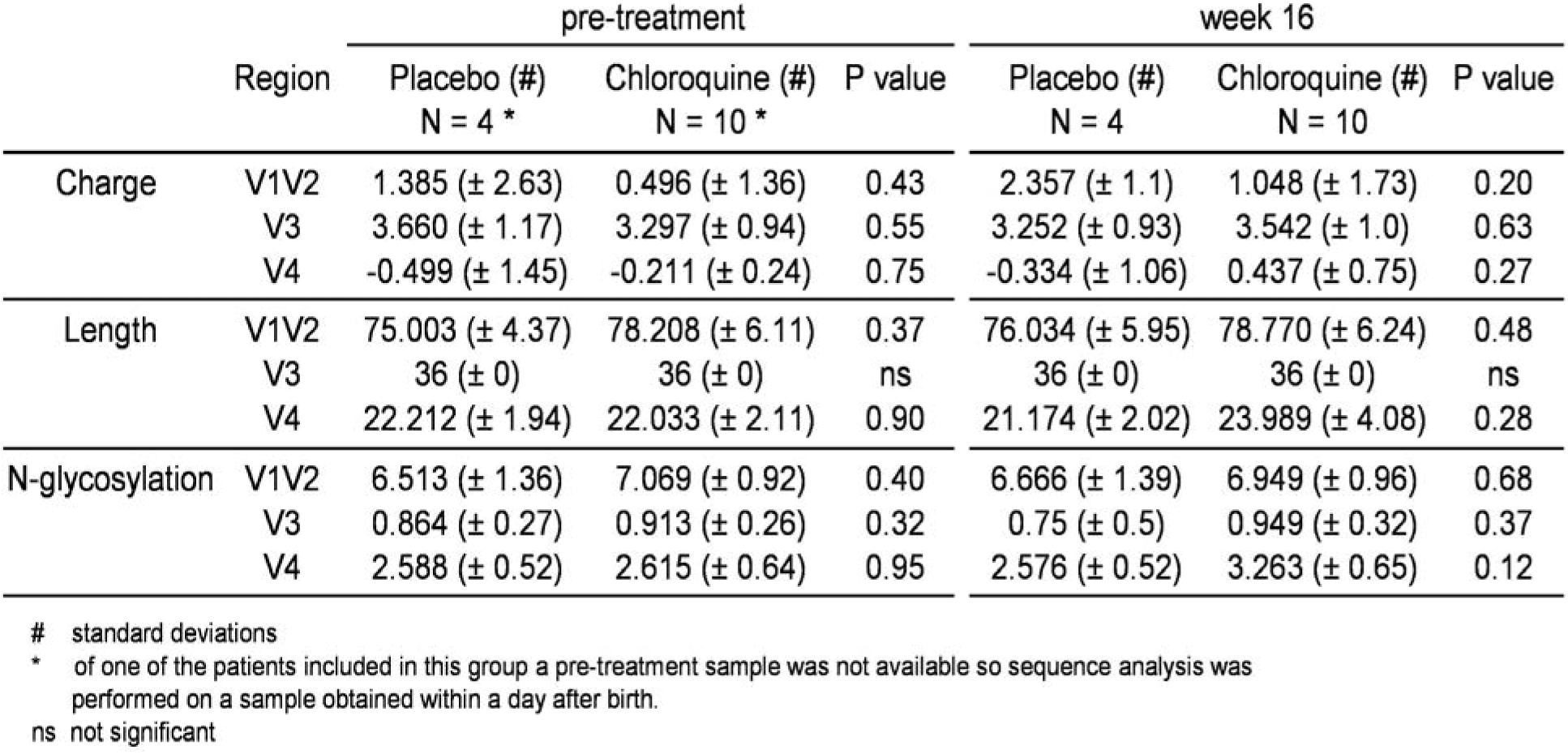
Sequence analysis of gp120 of CQ and Placebo treated group

**Fig. 1.**
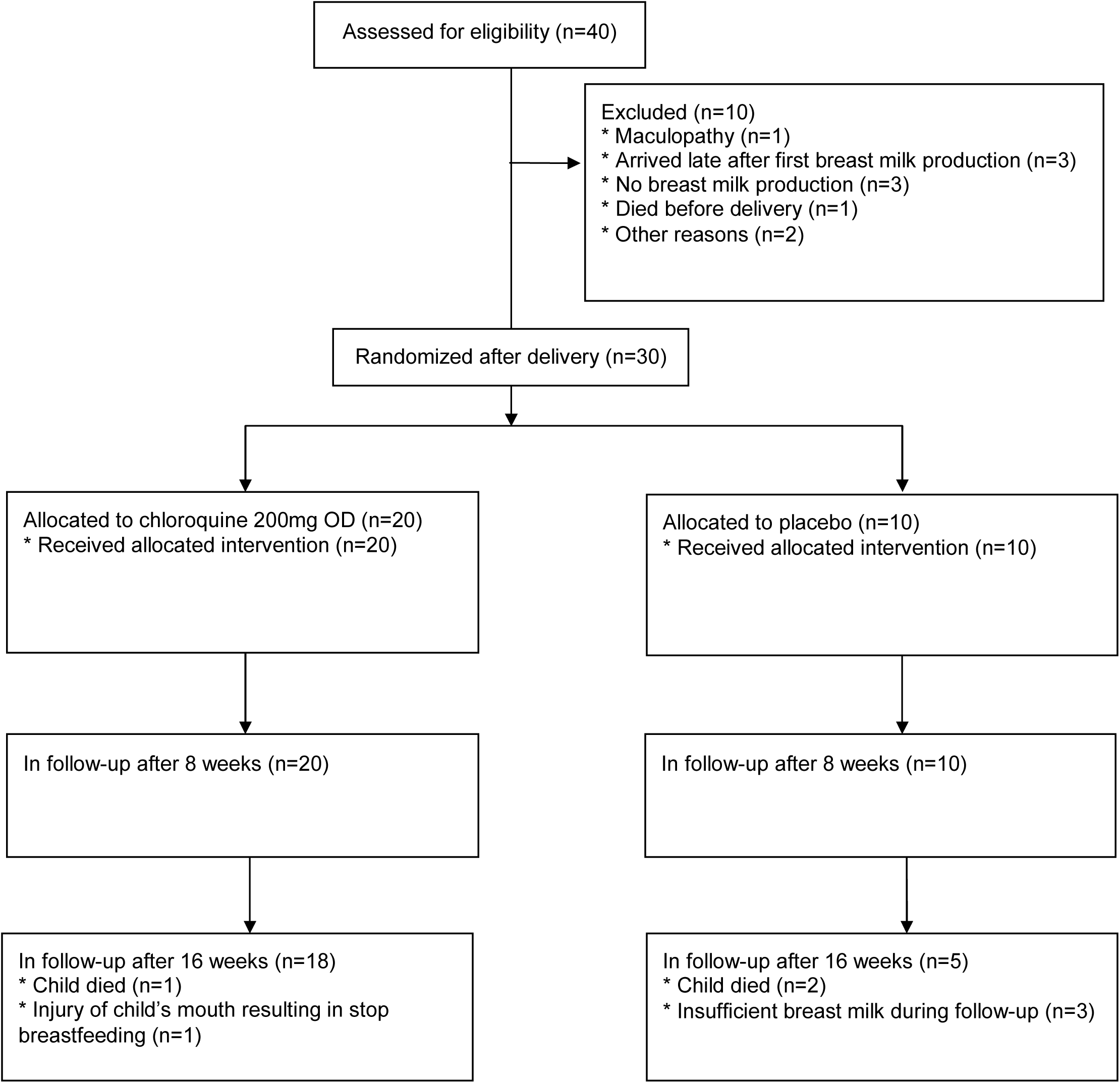
Schematic of the proposal study design. 30 mothers were enrolled with 20 belonging to the treatment arm and 10 to the placebo group.

### CQ concentrations in breast milk are higher than in plasma

No differences in mean CD4 counts were identified between the Placebo and CQ groups at either wk0 or wk16, or between the different wks for both groups. The mean CQ concentration in plasma at wk8 is 759.7ng/ml (244–877ng/ml) and at wk16 478.2ng/ml (174– 414ng/ml). Of note, there was an association between high plasma CQ concentrations at wk8 with high CD4 counts, with those harboring concentrations of CQ over 1,200 ng/ml having the highest CD4 counts (*P* < 0.0001) but which did not correlate with pVL at wk16 (data not shown). CQ levels in breast milk are higher than in plasma at both wk8 and wk16 (Fig. 2a) with a positive correlation between CQ levels in plasma and breast milk at wk8 and wk16 (Fig. 2b). In summary, CQ levels are higher in breast milk than in plasma and at wk8 the CQ levels are on average 2.5 fold higher in breast milk.

**Fig. 2.**
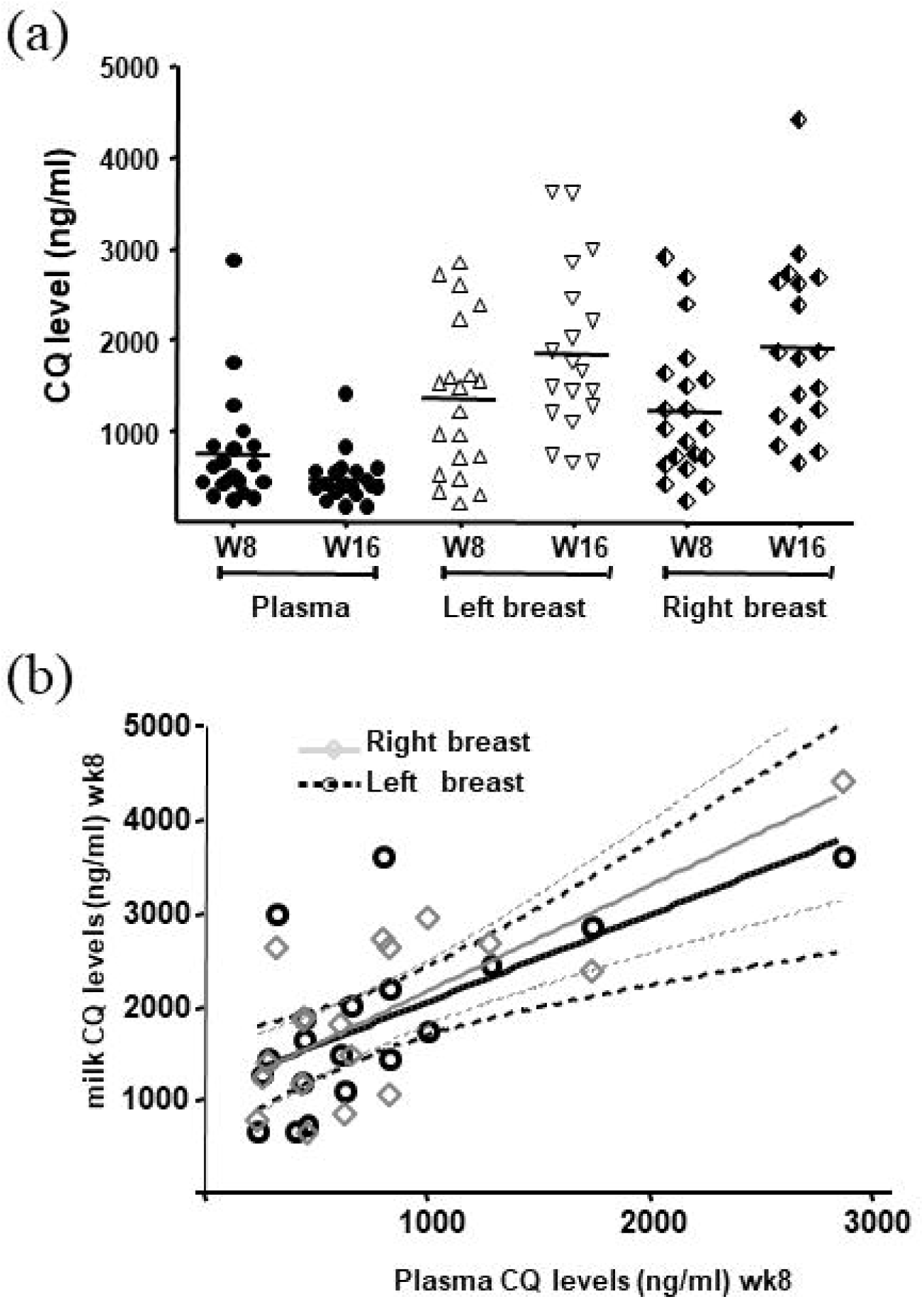
CQ levels after treatment. (a) CQ levels in the plasma, left breast and right breast of each study individual at wk8 and wk16 of treatment, with mean values depict. (b) correlation between plasma CQ levels and milk CQ levels at wk8

### CQ treatment results in increased plasma but not breast milk HIV-1 viral loads

Mean pVL for CQ and Placebo groups are shown in Fig. 3a and 3c, respectively. No differences were observed between psc and wk0 or wk8 and wk16 for either group. The pVL values were, however, higher in the Placebo group compared to the CQ group at both psc and wk0 (2.5- and 2.1-fold, respectively). When comparing pVL measurements before and after treatment we observed a ***significant increase*** in the CQ group between psc and either wk8 or wk16 (*P* < 0.001) as well as wk0 and the later time-points (*P* < 0.001) (Fig. 3b). The mean pVL in the CQ treated group shows an increase of 1.0 log after wk8. Similar conclusions are drawn when we calculate the fold increase in pVL between wk8/wk0 or wk16/wk0, shown in Table 1 and Fig. 3b. The results show that over half of the CQ treated individuals harbor large increases in pVL by wk8, with the others showing little changes. We found bmVL to be consistently lower than pVL with many below the detection limit and with no differences between the CQ and Placebo groups (Fig 3a and 3c, respectively). When we compare the pVL with the bmVL in the CQ group we find a positive correlation (P<0.05). We also observe no difference in bmVL fold increase when comparing wk8 or wk16 with psc or wk0 for either the CQ or Pl group (Fig. 3b and Fig. 3d, respectively).

**Fig. 3.**
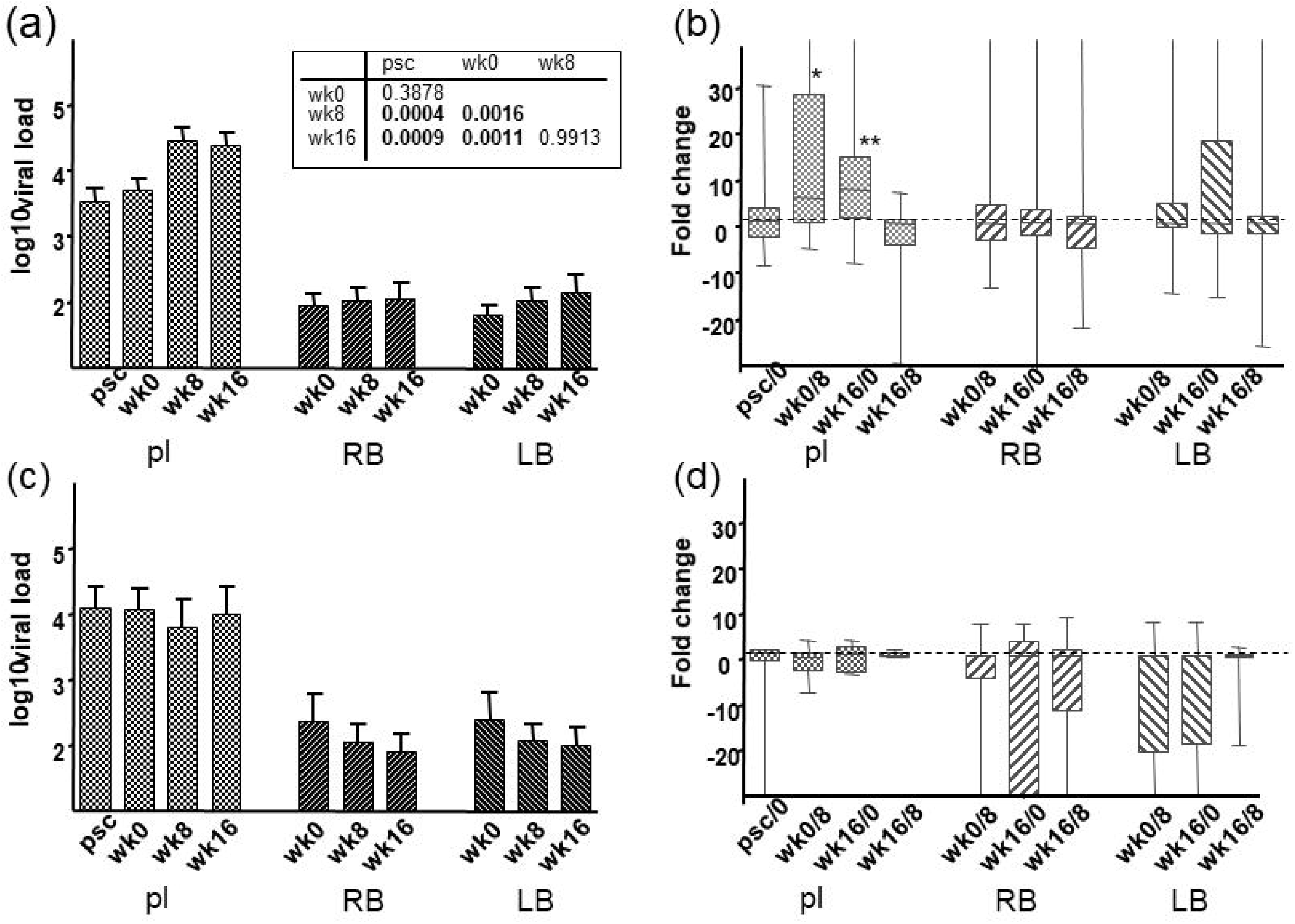
HIV-1 viral load measurements before and after CQ treatment. (a) mean HIV-1 viral load measurements for plasma, LB and RB at psc, wk0, 8 and 16 for the CQ treated individuals (b) fold increase in viral load measurements in plasma, LB and RB between different weeks for the CQ treated individuals (* *P* < 0.05, ** *P* < 0.005) (c) mean HIV-1 viral load measurements for plasma, LB and RB at psc, wk0, 8 and 16 for the Pl treated individuals (d) fold increase in viral load measurements in plasma, LB and RB between the different weeks for the Pl treated individuals.

### gp120 envelope genetic analysis before and after CQ treatment

Since we have previously reported genetic changes in the envelope of viruses passaged in the presence of CQ [6], we analyzed the gp120 C1C4 region DNA sequences in plasma from Pl (n = 4) and CQ individuals (n = 10) as designated in Table 1, before and after treatment (wk16). When comparing the overall amino acid charge of the V1V2 region we identified an increase over time for both the Pl and CQ treated groups and for the CQ treated individuals we found a link with increased pVL (*P* = 0.028), which was not observed for the Pl group. The V3 charge, which has been linked to coreceptor usage [21,22], was found to increase in 5/10 of the CQ treated individuals versus 1/4 of the Pl group, but was not statistically significant nor could be linked to increased pVL. A similar observation was found in the V4 regions. When comparing the alterations to the length of the variable regions no differences were found for either the Pl or CQ treated groups or with higher pVL in the CQ group. Similarly, alterations in the number of PNG sites did not alter in any of the variable regions in either the Placebo or CQ groups or between wk0 and wk16 of CQ treated individuals where pVL increased. Collectively, the results indicate no strong selection pressure exerted by CQ during the 16-week study period.

## Discussion

High levels of CQ could accumulate in both plasma and breast milk, with substantially higher levels accumulating in breast milk. CQ treatment did not correlate with reduced HIV-1 bmVL, but surprisingly, did associate with increased pVL at both wk8 and wk16. This is in stark contrast to previous reports which have described decreases in pVL after CQ treatment [3,4]. Given the CQ concentration administered and a previous study showing that the drug dosage prescribed correlates to HCQ blood levels [23] it is unlikely that we have reached insufficient concentrations of the drug. Although the majority of *in vitro* studies to-date have described lower replication of HIV-1 in the presence of CQ, one study has indicated that high concentrations of CQ can be associated with increased viral entry [24]. It should also be noted that in animal models for various virus infections, including Semliki Forest Virus (SFV), CQ treatment can result in increased viral replication and modulate cytokine responses *in vivo* [25].

It has been reported that CQ treatment can decrease the glycosylation pattern of the HIV-1 gp120 protein [1]. When we compare the alterations to gp120 sequences from before and after CQ treatment we did not observe any major alteration to the V1-V2, V3 or V4 sequences, suggesting that CQ had no effect on selecting variants with altered replication phenotypes. Although glycosylation differences were not observed at the amino acid level it does not rule out the possibility that post-translational modification differences account for the results obtained i.e. altered recognition of HIV-1 by the neutralizing antibody repertoire leading to immune escape.

CD8^+^ T-lymphocyte responses have been shown to be modulated by CQ treatment, with higher levels of cellular activation mediated through an increase in antigen presentation by MHC class-I molecules [26]. The same scenario may hold true for CD4^+^ T-lymphocytes with an increase in antigenic presentation via MHC class-II molecules by antigen presenting cells, resulting in heightened cellular activation and increased viral replication. This, however, would not explain the lack of increase in pVL in previous CQ or HCQ studies [3,4]. The present results support a recent study where mothers receiving CQ did not show a decrease in pVL or bmVL [27]. The previous studies with CQ and HCQ have all been performed on male participants and we cannot rule out gender effects or differences due to child birth and/or lactation. It has been shown that bacterial translocation from the gut can be associated with heightened immune activation in HIV-1 infected individuals [28]. Whether CQ can influence bacterial growth in the gut or at the vaginal mucosa and ultimately modulate immune activation in treated mothers soon after delivery needs to be studied. It has been suggested CQ can restrict signaling via the toll like receptor 9 (TLR9), however, it has also been shown that with intestinal epithelial cells, CQ can augment cellular activation and thereby possibly upsetting colonic homeostasis [29].

It has been hypothesized that CQ could be used as a safe and cheap alternative to conventional HIV-1 antiretroviral pMTCT regimes and has been considered for use in resource-limited settings. More recently chloroquine as well as hydroxychloroquine treatment has been advocated for inhibiting SARS-CoV-2 replication and treating COVID-19 patients. Our results indicate caution when treating HIV-1 positive women in the post-natal period with CQ and especially those soon after childbirth where the likelihood of HIV-1 transmission through breastfeeding is present. Increased HIV-1 viral loads in mothers receiving CQ, opposed to the drugs observed inhibitory effects *in vitro*, provide caution in prescribing the drug as an effective antiviral to all patients. Increased viral loads in plasma of post-partum mothers may reflect hormonal effects which have been shown to associate with modulation of immune responses and where CQ treatment may have an effect on influencing HIV-1 activation and replication [30]. It has also been documented that CQ can have differential effects on activation of transcription factors, such as through NF-κB, with the potential to induce pro-inflammatory responses [31], with the potential consequences of increasing HIV-1 replication and plasma viral loads.

Several commentaries have proposed using CQ and HCQ to treat coronavirus COVID-19 as part of a ‘drug repositioning’ strategy [32–39]. CQ has displayed in vitro activity against the SARS-CoV-1 [40–43]. Middle East respiratory syndrome coronavirus (MERS-CoV) [44], human coronavirus 229E (HCoV-229E) [45] and HCoV-O43 [46,47]. While both HCQ and CQ display in vitro activity against SARS-CoV-2 [48,49], the evidence proposes HCQ as potentially exhibiting a stronger in vitro anti-SARS-CoV-2 activity, of the two drugs. The in vivo evidence is less distinct in that a study treating confirmed coronavirus patients with HCQ sulphate outperformed the control group ^[23]^ whereas a Shanghai ^[25]^ study reports the control having a slightly better outcome. It is apparent that more studies are required to inform the use of CQ or HCQ for SARS-CoV-2 treatment. Our findings of increased HIV-1 pVL with CQ in post-partum women should be taken into consideration when designing or administering CQ or HCQ in COVID-19 clinical trials, especially relating to the post-partum setting.

## Data Availability

All data will be made available

## Acknowledgements

This work was supported through Dutch AIDS Funds (6013) and Elizabeth Glaser Pediatric AIDS Foundation (27-PG-51269). We thank Dr Michel de Baar for critical review of the manuscript and we are indebted to the mothers and infants who participated in this study at the CHUK.

## Author Contributions

SL, JV, RWS, JHB, JRB, WAP and JML were involved with the clinical and scientific design of the study. SL was the Principal Investigator overseeing the study. SL, JV, FWW, NJV, BK and RD were involved with the clinical implementation of the study, data collection and clinical data analysis. MAN, RWS, SJ, JHB, GP and WAP were involved with experimental design and scientific data analysis. MAN, ST, performed experiments. WAP, MAN, MC, SL, wrote the manuscript.

## Notes

### Competing Interest Statement

The authors have declared no competing interest.

### Clinical Trial

STEG-METC (ref no R01-089)

